# Olfactory Dysfunction in the COVID-19 Era: An Umbrella Review Focused on Neuroimaging, Management, and Follow-up

**DOI:** 10.1101/2023.02.07.23285588

**Authors:** Mohammadreza Kalantarhormozi, Houman Sotoudeh, Mohammad Amin Habibi, Mehdi Mahmudpour, Ramin Shahidi, Fattaneh Khalaj, Shaghayegh Karami, Ali Asgarzadeh, Mansoureh Baradaran, Fatemeh Chichagi, Sara Hassanzadeh, Narjes Sadat Farizani Gohari, Mahsa Shirforoush Sattari, Amir Azimi, Ali Dadjou, Mahsan Eskandari

## Abstract

**Purpose:** The coronavirus disease 2019 (COVID-19) is surrounded the world and is associated with multiorgan damage. Olfactory dysfunction is a common manifestation in COVID-19 patients, and in some cases, presents before the coryza signs. We conducted this umbrella review to provide a practical guide on managing, imaging findings, and follow-up of COVID-19 patients with olfactory dysfunction (OD).

**Methods:** A comprehensive search was performed in PubMed, Embase, Scopus, and Web of Science databases from December 2019 until the end of July 2022. Systematic reviews and meta-analyses addressing management and imaging findings of the olfactory manifestations of COVID-19 were included in the study. The quality assessment of included articles was carried out using the Assessment of Multiple Systematic Reviews-2 (AMSTAR-2) tool.

**Results:** A total of 23 systematic reviews were reviewed in this umbrella review. The number of included studies varied between 2 to 155 articles. Several demographic variables were not adequately reported across all the included systematic reviews, including age, gender, preexisting comorbidities, or whether participants had been hospitalized or admitted to the intensive care unit (ICU) due to COVIDLJ19.

**Conclusion:** It seems that the coronavirus can infect olfactory system structures that play roles in the transmission and interpretation of smell sense. Based on studies, a large proportion of patients experienced OD following COVID-19 infection, and the majority of OD was resolved spontaneously. The possibility of long-lasting OD was higher in young adults with moderate clinical manifestation. Olfactory training (OT) was the most effective therapy. Intranasal corticosteroids (ICS) are also recommended.

## 1. Introduction

The novel severe acute respiratory syndrome coronavirus 2 (SARS-Cov-2) virus was a siege on the world and imposed a massive burden on the healthcare system [1]. The disease (COVID-19) caused by this virus can involve multiple organs [2]. Involvement of the olfactory system and OD can affect patients’ quality of life [3, 4]. Loss of smell is a well-known clinical manifestation of COVID-19-infected patients and can be associated with direct and indirect complications such as poisoning [5] and psychological and cognitive disorders [6]. OD has been reported to involve between 22% and 85.6% of COVID-19 patients [7, 8].

The OD sustains in 75% of the patients with persistent COVID-19 manifestations (Long COVID) [9]. Given the COVID-19 pandemic and the high prevalence of OD in this disease, otolaryngologists encounter challenges in diagnosing, treating, and following up on COVID-19 patients with OD.

Currently, the data related to the diagnosis, management, and follow-up of COVID-19 patients with OD is disputable, while definitive evidence is required to treat these patients. Here, we sought to provide accurate and reliable evidence regarding the challenging aspects of managing COVID-19 patients with OD. To address these issues, in this review, we directed a comprehensive umbrella review on the systematic reviews (SR)/systematic reviews and meta-analysis (SRMA) to give definitive information regarding the imaging findings, management, and follow-up of COVID-19 patients with OD and reduce the dilemmas.

## 2. Methods and materials

The present umbrella review was completed according to Preferred Reporting Items for Systematic Reviews and Metaanalysis (PRISMA) [10] and the guideline of umbrella review designed by Aromataris et al [11].

### 2.1. Search strategy

A comprehensive literature review was performed on 31 July 2022 to identify systematic reviews and meta-analysis, which provided data on the neuroimaging, management, and follow-up of olfactory manifestation of COVID-19. PubMed, Embase, Scopus, and Web of Science databases were systematically searched from December 2019 until the end of July 2022 using related keywords that were extracted from terms in related articles. These terms were as follow: “COVID-19”, “coronavirus disease 2019”, “SARS-CoV-2”, “severe acute respiratory syndrome coronavirus 2”, “olfactory”, “systematic review”, and “meta-analysis”. A search strategy was designed for each database by combining these keywords with appropriate Boolean operators (OR/AND) and searching within the titles, abstracts, Medical Subject Headings (MeSH), Emtree, and other search fields. These strategies are summarized in table S1 (supplementary materials). Results of the systematic search were complemented by the manual addition of eligible articles found by checking reference lists of primary selected studies in the Google Scholar database.

### 2.2. Eligibility criteria

All of the peer-reviewed systematic reviews, with or without meta-analysis that addressed the treatment, neuroimaging, and follow-up of OD in COVID-19 patients were included. PICO items are defined as follows:

**P (Population):** COVID-19 patients who affected by OD

**I (Intervention):** Treatment with different approaches such as OT

**C (Comparator):** COVID-19 patients who are not affected by OD

**O (Outcome):** Recovery from OD or persistent symptoms

Exclusion criteria were: (1) the other types of studies include primary articles (experimental, cross-sectional, interventional, and longitudinal studies), narrative reviews, and grey literature, (2) studies with no report on the OD manifestations of COVID-19, (3) studies focusing on other viral diseases, (4) studies without available English full-text, (5) The pre-prints and not peer-reviewed publications also were excluded.

### 2.3. Study selection process

The search findings were transferred into EndNote X9, and duplicates were removed. Two reviewers (A.As and MA.H), independently selected potentially relevant studies based on their title and abstracts. Then, the same reviewers conducted the full-text assessment of studies for the final inclusion. The disagreements were dissolved by referring to a third reviewer (M.E).

### 2.4. Data extraction

Two independent reviewers (M. SS, F.K, and A.Az) investigated the full texts of the included articles and the following data were collected by using an Excel worksheet: first author’s name, year of publication, searched databases, study designs (systematic review and/or meta-analysis), number of the included studies, Number of patients, duration of symptoms, type of manifestations, and main findings. The extracted data were reassessed through a third investigator (SH. K) and the missed data was added to the worksheet.

### 2.5. Quality assessment

Two reviewers (M. SS, F.K, and A.Az) independently evaluated the quality of the comprised articles with the Assessment of Multiple Systematic Reviews-2 (AMSTAR-2) tool [12]. They categorized the quality of reviews into critically low, low, moderate, or high. Then, a third reviewer (SH. K) rechecked the included articles’ quality assessment to ensure the studies’ scoring.

### 2.6. Data synthesis

A qualitative review of the included studies was carried out and their characteristics were summarized in the tables below. Statistical analysis was not performed due to the heterogeneity of the systematic reviews and missing data about the overall effect sizes of each outcome.

## 3. Results

### 3.1 Study selection

After removing the identical studies from a total of 344 studies, 102 articles remained for screening the titles and abstracts. Twelve irrelevant articles were removed and 90 studies were investigated in full-text assessment. For further analysis, 23 manuscripts met all criteria of this umbrella review. Figure 1 shows the PRISMA flowchart to summarize the process. The quality assessment results based on AMSTAR-2 are presented in Table S2 (supplementary materials).

**Figure 1.**
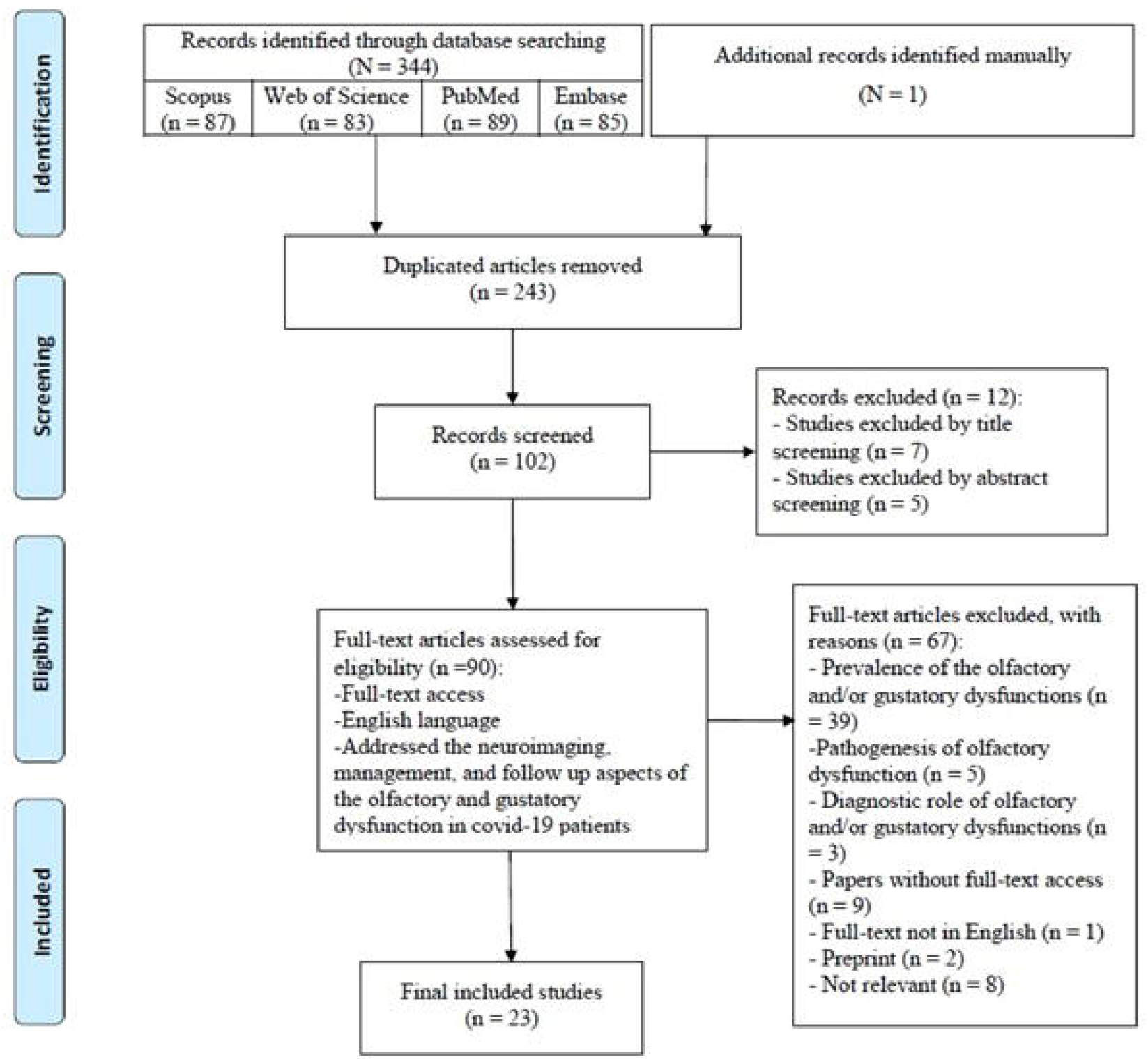
PRISMA Flowchart

### 3.2 Characteristics of included studies

This umbrella review included 23 systematic reviews (and Meta-Analysis) published in English from inception to July 2022. All of these publications used different databases to retrieve primary studies. The number of recruited electronic databases varied from 1 to 10, whereas MEDLINE and EMBASE databases are the dominant information sources across all reviews. The number of primarily included studies in different reviews varies from 2 to 155. Several demographic variables were not adequately reported across all the included systematic reviews, including age, gender, preexisting comorbidities, or whether participants had been hospitalized or admitted to the intensive care unit (ICU) due to COVID□19. Incomplete data present a limitation since all of these factors may impact the post-COVID experience. The length of olfactory dysfunction (OD) varies from 1 week to 6 months through different reviews. Studies have generally reported hyposmia and anosmia as the predominant symptoms of OD, whereas parosmia and Phantosmia are less frequently reported.

### 3.3 Studies on neuro-imaging findings

Eight articles assessed the neuroimaging findings of COVID-19 patients with OD. Several neuroimaging modalities were applied to delineate the impression of COVID-19 on olfactory systems, including computed tomography (CT) scan, positron emission tomography (PET), and magnetic resonance imaging (MRI) [13-19]. Moreover, different Structures were evaluated to highlight the impact of COVID-19 on the OD, such as Olfactory cleft, olfactory bulb, olfactory tract, olfactory sulcus, olfactory gyrus, and brain cortex related to olfactory sensation. Also, figures 2-5 illustrate the different types of involvement of the olfactory system in MRI and CT studies.

**Figure 2.**
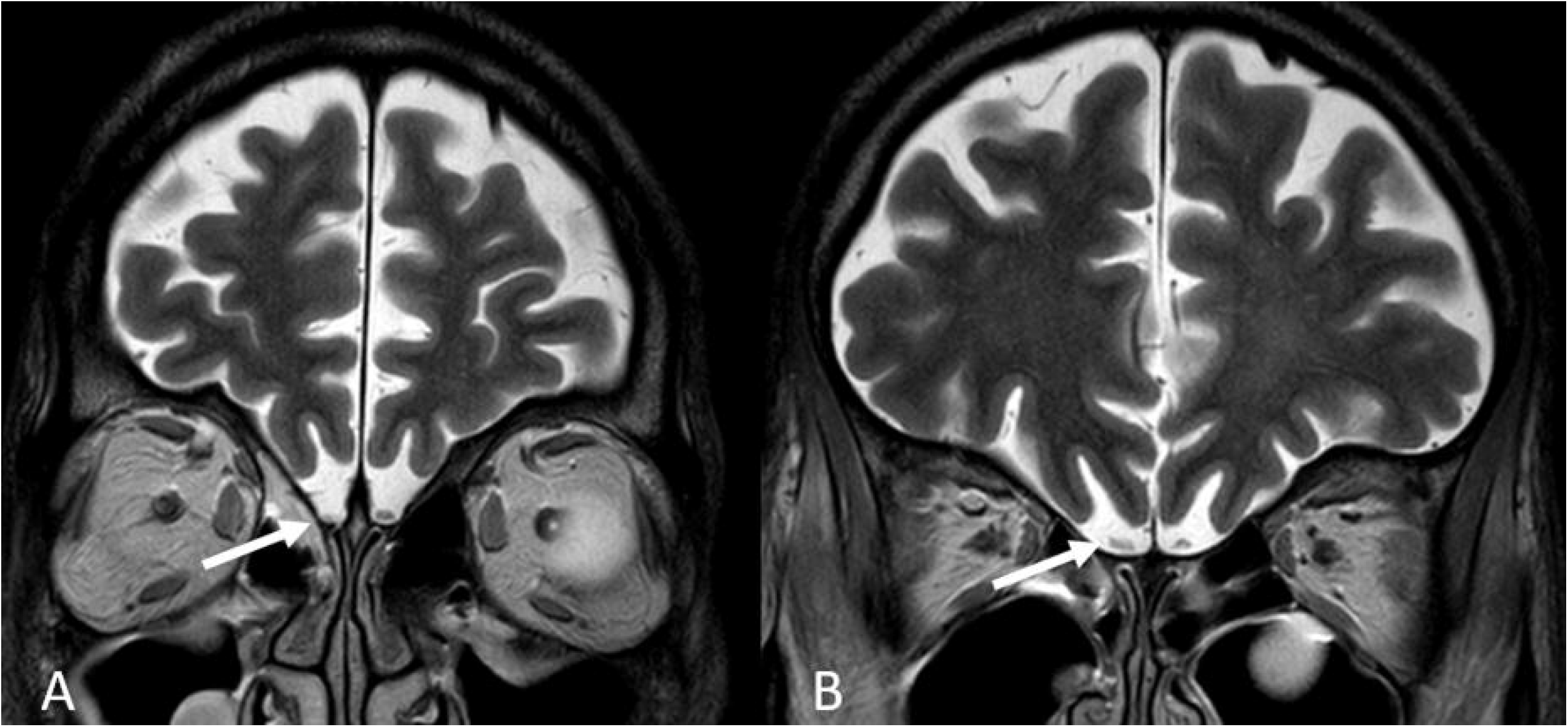
51-year-old man with COVID-19 infection. CT shows Opacification of the left olfactory cleft in different views, arrows (**A**, axial CT, **B**, Coronal view).

**Figure 3.**
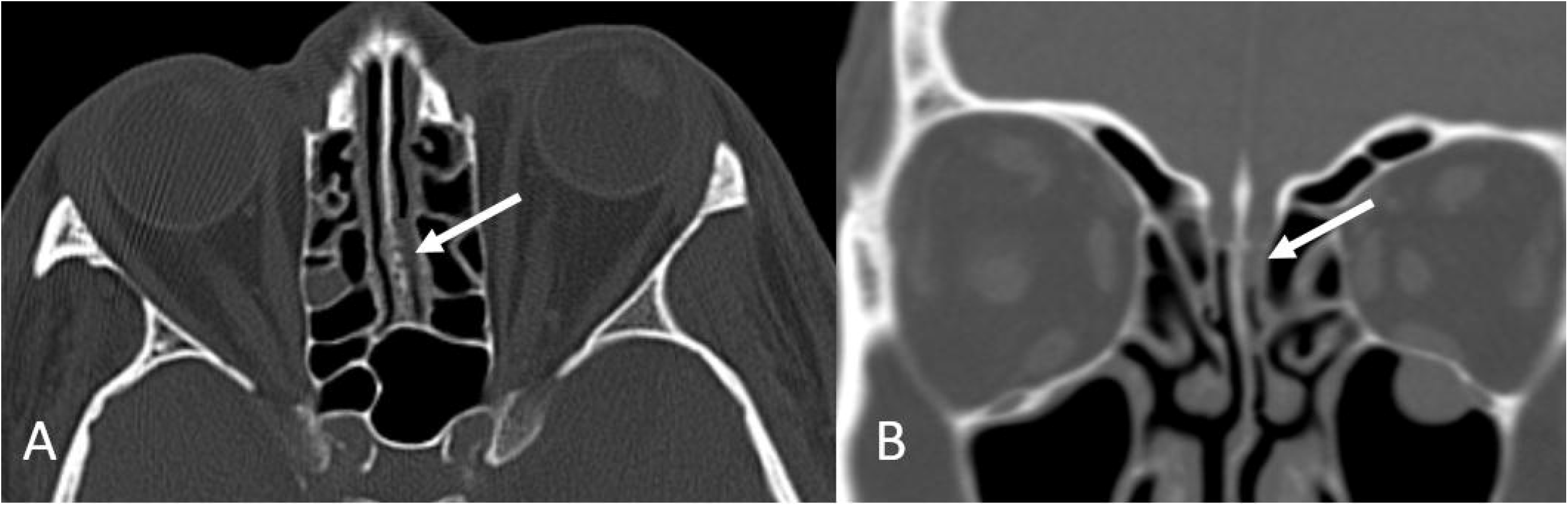
60-year-old man with COVID-19 infection. Coronal T2 sequence shows T2 hypersignal intensity of the right olfactory bulb (A and B, arrow). Swelling of the right olfactory bulb is seen more posteriorly (B, arrow).

**Figure 4.**
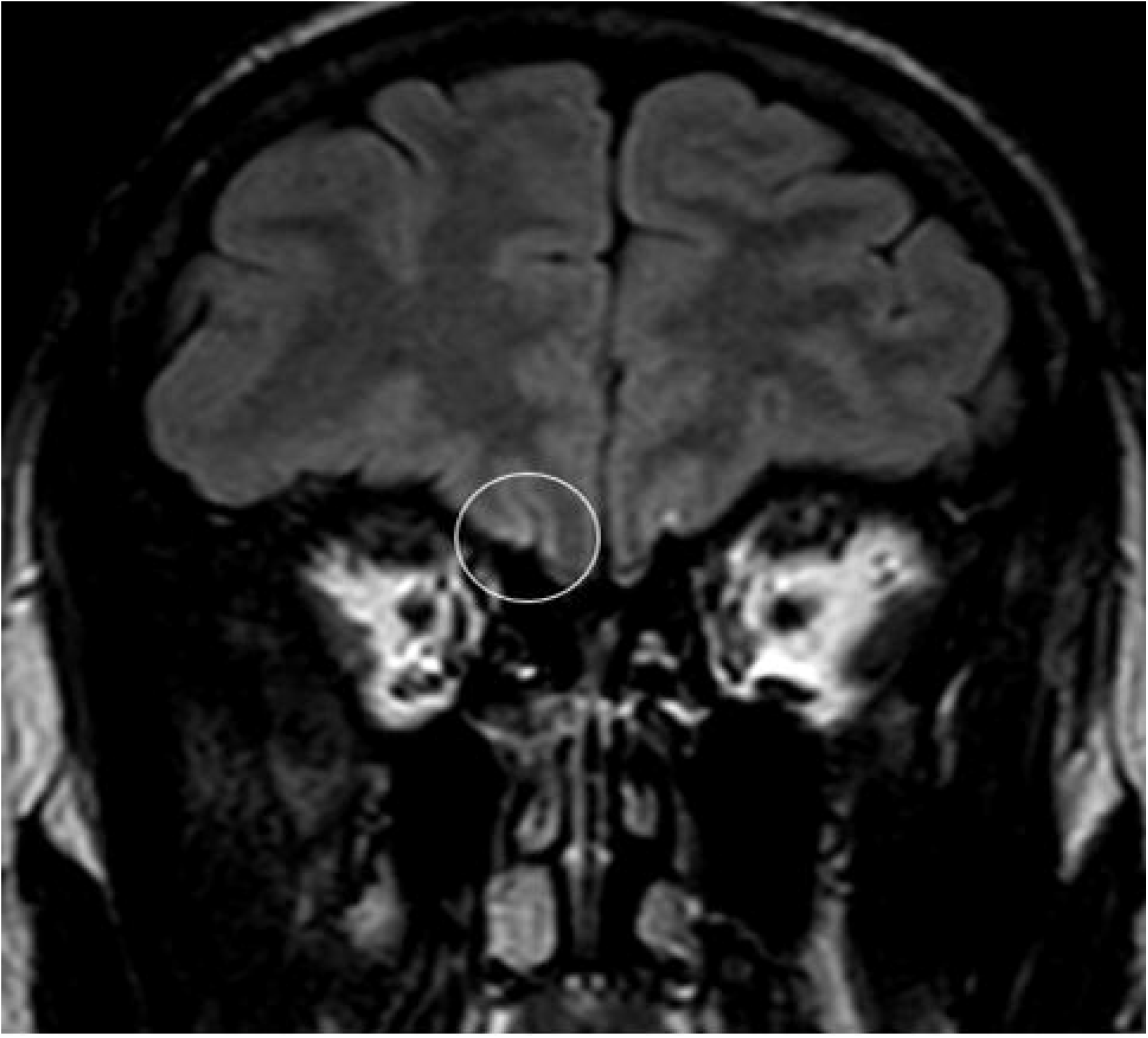
19-year-old man with COVID-19 infection. Coronal FLAIR sequence shows subtle sulci FLAIR hypersignal intensity of right olfactory sulcus (marker).

**Figure 5.**
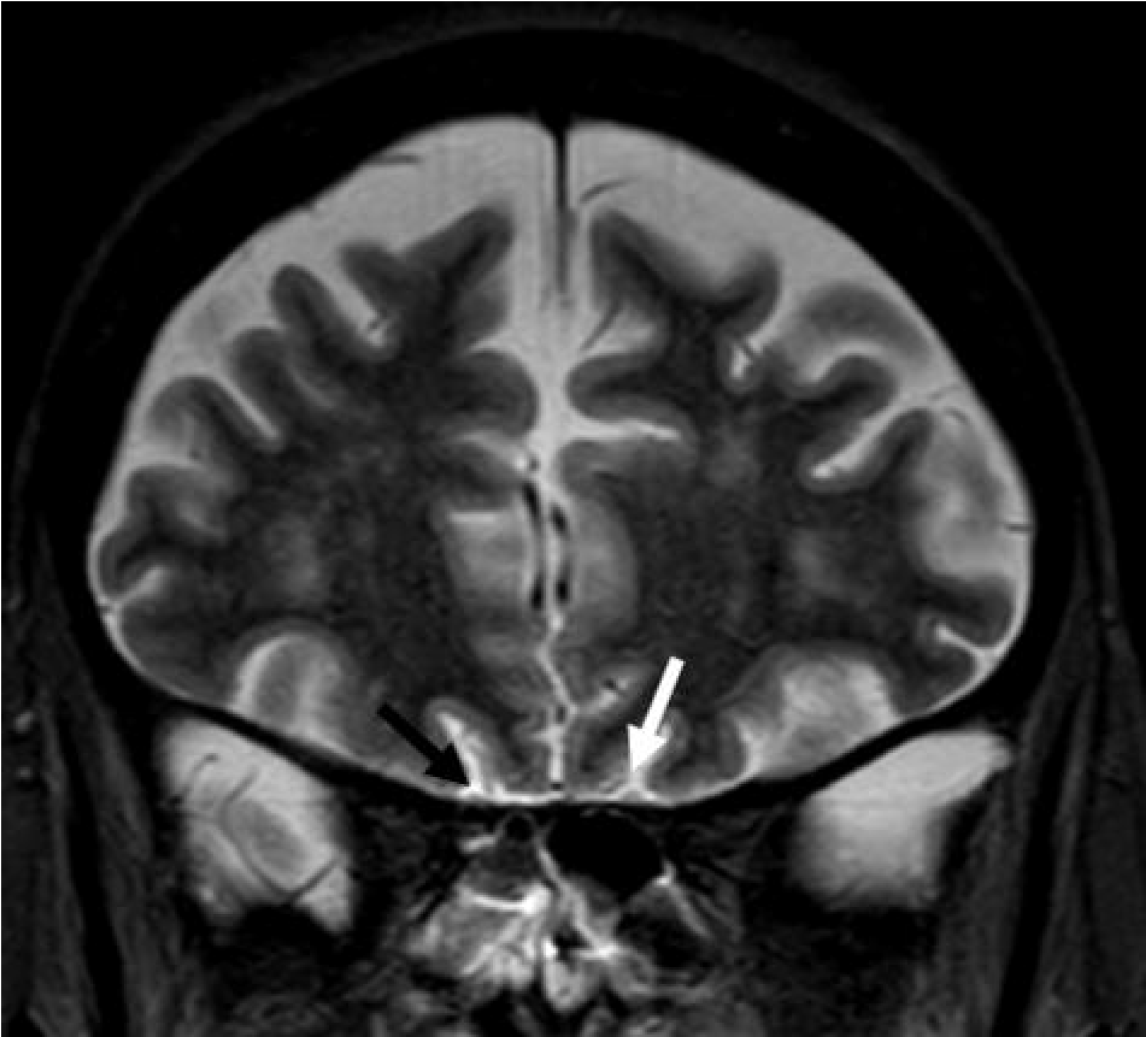
62-year-old woman with COVID-19 infection. Coronal T2 sequence shows subtle edema of left olfactory tract (white arrow) in comparison to right side (black arrow).

#### 3.3.1 Olfactory cleft (OC)

The imaging findings of olfactory cleft (OC) in COVID-19 patients were reported in seven reviews [13-19]. In the visual inspection of the MRI images, inflammation and obliteration of the OC were observed, suggesting that the virus has reached the brain through the olfactory nerves [14]. Complete or incomplete obstruction of the OCs with mucosal thickening on CT [13] and mucosal thickening with T2 hyper signal intensity on MRI are among the reported pathologies of OC in COVID-19 [16, 19].

Another manifestation reported in two reviews is an increase in the width and volume of OC. Enlargement of the OC increased T2 signal intensity [19], and the widening of OC [18] have been reported. An important radiological marker of COVID-19-associated OD is the presence of opacification in the olfactory cleft on CT or MRI imaging. Compared to control subjects, patients with COVID-19 infection and olfactory dysfunction (OD) had significantly higher levels of olfactory cleft opacification [17].

#### 3.3.2 Olfactory bulb (OB)

Six reviews described the imaging findings of olfactory bulbs (OB) in COVID-19 patients [13, 15-19]. It was demonstrated that no significant differences were observed between COVID-19 patients with OD and non-COVID-19 individuals in terms of mean volume of OB [18]. However, the altered dimension of OB is a common finding in other studies. For example, considerable enlargement and edema of bilateral OB were reported seven days after the diagnosis of COVID-19 [17]. Conversely, a decreased volume of OB characterized by asymmetry, thinning, and loss of standard oval shape is reported more commonly in the later stage of infection [13]. In most cases, OB appears normal within the first few days following olfactory dysfunction, but deformation and degeneration of OB are more evident in the delayed phase and are associated with longer chemosensory impairment [13, 15, 19]. Therefore, it appears that the timing of the imaging during COVID-19-induced olfactory dysfunction is essential in detecting abnormalities in OB imaging.

The most frequent overall MRI result was normal OB morphology and signal in the early and late imaging [13]. However, it was reported that approximately half of the patients have an abnormal olfactory bulb morphology on MRI with mild irregularity of the olfactory bulb by a preserved J-shape, contour lobulations, and rectangular-shaped olfactory bulb [17].

In addition, altered olfactory bulb signal intensity was frequently described, with diffuse hypointense foci resembling microhemorrhages, abnormal T2 FLAIR signals, and olfactory bulb inflammation with abnormal enhancement demonstrated by pre and post-contrast fat suppression images [13, 16, 18, 19]. Although, one study has stated that patients and controls seem to have an equal proportion of olfactory bulb signal abnormalities [17].

#### 3.3.3 Olfactory sulcus, gyrus, and cortex

Brain imaging findings of the olfactory sulcus, gyrus, and cortex in individuals with COVID-19 and OD were described in five studies [13, 14, 17-19]. Cortical areas responsible for processing olfactory stimuli were shown to be altered. Gyrus rectus changes as hyperintensity on FLAIR MRI, and significantly lower metabolic rate in right rectal gyrus on 18F-FDG brain PET imaging [19], bilateral cortical hyperintensity involving the rectus gyrus and hypometabolism of bilateral olfactory rectus gyri on 18F-FDG PET [17], hyperintensity in the orbitofrontal and entorhinal cortices of the right rectus gyrus and the left caudate and para-hippocampus on early and late imaging, T2 hyperintensity and CT hypodensity in the cortex of bilateral inferior frontal lobes and the straight gyrus [13], and changes in the olfactory area as well as adjacent brain regions, including the limbic and prefrontal cortex, with the extension of the abnormalities to the olfactory brain network, which is implicated in the corpus callosum, cingulate cortex, and insula and altered volume, thickness, and hypometabolism in the olfactory cortex have been reported [14]. On the other hand, one study found no statistically significant difference in the right olfactory sulcus depth between COVID-19 patients and non-COVID-19 controls [18].

#### 3.3.4 Other olfactory structures

Several reports have been made concerning other olfactory structures, such as the olfactory nerves, tracts, and paranasal sinuses. Three reviews demonstrate olfactory tract abnormalities [17-19] with evidence of bilateral T2-FLAIR hyperintensities, enhancement, and DWI abnormalities indicative of olfactory tract inflammation [19], and enlargement of the central olfactory tract, which is more prominent on the right side[17]. In addition, a significant reduction in the mean length of right and left olfactory tracts in patients with COVID-19 and persistent OD in comparison to normosmic healthy controls [18] has been noted in these studies. One study revealed destruction in the olfactory nerve morphology and filia architecture by the clumping and thickness of the olfactory filia, as well as a scarcity of them on olfactory nerve MRI [17]. The manifestation of paranasal sinuses is discussed in two articles [13, 17]. MRI demonstrated minor mucosal thickness in the ethmoid sinuses and inflammation or partial obliteration in the ethmoid, sphenoid, and/or maxillary sinuses [17]. However, in another study, the paranasal sinuses were found to be normal in most of these scans [13].

### 3.4 Studies on the management of COVID-19-related OD

#### 3.4.1 Steroids

Some studies have examined the use of steroids in treating patients with post-infectious olfactory dysfunction (PIOD) using various formulations, routes, and doses of steroids [20-23]. An oral steroid trial for a limited period can resolve residual mucosal congestion related to the PIOD and improve the therapeutic effects of other techniques (smell training, spontaneous recovery). It is well-recognized that topical steroids have fewer side effects than systemic steroids [24]. Nevertheless, administration instructions should be considered carefully, as nasal steroids are most effective when sprayed via nozzles (such as laryngeal mucosal atomization devices) or as nasal drops in a dependent head position. [20]. Treatment with topical mometasone has been shown to improve odor threshold and identification [22]. The other review reported that topical steroid significantly affects the early phase of PIOD within four weeks. However, full recovery of PIOD is not achievable, and no significant difference was observed between patients using topical steroids and the placebo groups over four weeks after treatment [23]. One study assessed the combined effect of systemic steroid and steroid irrigation and showed the improvement of smell sense. However, recovery and changes in olfactory function, and adverse reactions to steroids remain uncertain [21].

#### 3.4.2 Olfactory training

In classic olfactory training (OT), four different odorants are exposed twice daily for five minutes [25]. Previous studies have found that these four odorants (phenyl ethyl alcohol, eucalyptol, citronella, and eugenol) improve olfactory loss of post-viral olfactory dysfunction after 12 weeks of training. Olfactory training is often recommended as a treatment that is relatively safe without adverse effects and may be utilized combined with other therapies. It was found that OT was effective in identifying and discriminating odors. Furthermore, MRI studies showed alterations in connectivity between cortical olfactory networks after OT. Subsequently, the OT has been recommended as the first line for treating PIOD [20, 22]. In one study, the use of OT in combination with steroids was compared to olfactory training alone and showed that adding the nasal steroid spray can not provide any additional benefit [26]. In the modified OT, the four odorants are initially used for 12 weeks, the same as in classic OT. In the following 12 weeks, the second set of odorants (menthol, thyme, tangerine, and jasmine) is applied, and then a third round of odorants (green tea, bergamot, rosemary, and gardenia) for the final 12 weeks [20]. Recent data shows that this technique over 24 to 36 weeks is better in odor discrimination and identification than classical training.

#### 3.4.3 Other treatments

Some studies have investigated other therapeutic options for improving olfactory functions and reducing recovery time. The therapeutic agent recruited in different studies were summarized:

- *Zinc supplements:* Patients with PIOD treated with zinc sulfate have not demonstrated statistically significant improvements in their olfactory function [20, 22]. However, one study documented that zinc therapy significantly shortened the recovery time of olfactory dysfunction (14).
- *Retinoic acid (Vitamin A):* Supplementing with 10,000 IU vitamin A did not demonstrate any benefit compared to placebo. However, the administration of 10,000 IU of topical vitamin A along with OT resulted in significant clinical improvement over OT alone [20, 22].
- *Probiotic strains:* One research examined the intranasal delivery of L. lactis W136 to COVID-19 patients within 96 hours of their diagnosis. Intranasal treatment for 14 days resulted in a decrease in symptoms’ intensity and olfactory impairment (15).
- *Theophylline:* Theophylline is a phosphodiesterase inhibitor and it has been shown to increase olfactory function when administered orally [20, 22].
- *Sodium Citrate:* Intranasal sodium citrate showed improved olfactory threshold and identification due to its capacity to bind calcium ions. Intranasal sodium citrate is believed to lower free mucosal calcium, hence decreasing negative feedback and boosting sensitivity to odorants [20, 22].
- *Minocycline:* The anti-apoptotic activity of minocycline may contribute to improving olfactory function. However, minocycline, did not significantly improved odor sensations compared to the placebo [20, 22].
- *N-Methyl D-aspartate antagonist:* By avoiding glutamatergic neurotoxicity, it is hypothesized that Caroverine, an N-methyl-D-aspartate (NMDA) antagonist, can aid in treating post-viral olfactory dysfunction. Caroverine enhanced odor thresholds in anosmic patients and odor recognition in both anosmic and hyposmic individuals with post-viral olfactory dysfunction [20, 22].
- *Topical Intranasal Insulin:* topical intranasal insulin has been studied and shown to increase odor sensitivity in 60% of patients, with modest increases in odor discrimination [22].
- *Alpha Lipoic acid:* alpha-Lipoic acid (ALA) is a fatty acid primarily used to treat diabetic neuropathy. It also possesses antioxidative and neuroprotective effects and increases the production of nerve growth factors, substance P, and neuropeptide Y. It was recognized that applying ALA results in a modest increase in olfactory function [20].

### 3.5 Studies on follow-up and outcomes in COVID-19-related OD patients

Ten studies examined the long-term course and recovery rate from OD [19, 26-34]. According to two studies, the majority of OD cases remained chronic, and most patients recovered incompletely within the first weeks after the elimination of other symptoms. Moreover, in those who recovered from OD, recovery happened mostly within two weeks of the onset of symptoms [19, 32]. In contrast, in the other three studies, the percentage of spontaneous recovery of olfactory symptoms was reported to be very high [27, 29, 31]. In most reviews, Age, gender, and medical comorbidities were predictors of post-COVID-19 olfactory recovery. It is hypothesized that individuals aged<40 years have more tissue injury resistance than older individuals, and the age of >70 years is a predictor of poor long-term recovery from COVID-19-related OD [28], On the other hand, another study found that young adults are at an increased risk of olfactory dysfunction [34]. In contrast to two studies demonstrating a higher recovery rate in women [26, 28], another review concluded that women were more likely to continue the olfactory dysfunction [30]. A less severe form of COVID-19 disease in non-hospitalized patients is associated with a greater prevalence of anosmia, and longer symptoms [26, 31, 33]. Two other studies have suggested that highly severe cases have a higher risk of developing long-lasting anosmia and experiences a persistent deficit [28, 32].

## 4 Discussion

Globally, COVID-19 has been the biggest challenge for health care systems since December 2019. Most patients with Covid-19 suffer from olfactory dysfunction, which is associated with high healthcare costs for governments. Although most olfactory dysfunctions in covid-19 patients spontaneously resolved, some patients with persistent manifestations of covid-19 did not recover spontaneously, highlighting the importance of considering the different clinical aspects of these patients. Furthermore, several systematic reviews and meta-analyses have been published assessing different aspects of COVID-19 patients with OD and showed some controversies in the care of these patients. Considering this, it is imperative to conduct a comprehensive review of the current scientific evidence for different aspects of this patient’s care. Based on our included studies, we can discuss these patients in the following aspects.

### 4.1 Neuroimaging findings in Olfactory dysfunction

A meta-analysis of 17 cohort studies presented that 9.7% of COVID-19 patients underwent neuroimaging which 35.5% had abnormalities associated with the SARS-Cov-2 virus. Moreover, it was shown that these abnormalities were more common in critically ill patients [16]. Several studies used different neuroimaging techniques to determine the details of OD in COVID-19 patients. Although the underlying mechanism of olfactory dysfunction (OD) is not completely understood, investigating the mechanism of action of the SARS-Cov-2 virus can help to understand the exact pathophysiology of OD. According to previous studies, the SARS-Cov-2 virus interacts with angiotensin-converting-enzyme 2 receptors (ACE-2) [35], which are highly expressed in the nasal epithelium. Therefore, damage to the neuroepithelial cells can cause inflammation, impairment of neurogenesis in olfactory neurons, and lead to OD [36-38]. Based on neuroimaging studies, OD is associated with nasal epithelium abnormalities. In patients aged > 60, the virus DNA has been detected in olfactory areas, which makes it possible to damage this area and cause olfactory symptoms. Moreover, inflammation has been found in the olfactory epithelium, which may be associated with underlying mechanism of the SARS-Cov-2 virus-induced OD [39].

Despite the abovementioned facts, Mohammadi et al. [40] conducted a meta-analysis on 180 COVID-19 patients and 308 healthy subjects. They found no statistically significant difference between the COVID-19 patients and non-COVID-19 controls regarding the anatomic imaging of the olfactory system, including OB volume and OS depth.

Furthermore, pooled results of a Systematic review on case series, case-control studies, and case reports showed that olfactory cleft (OC) opacifications and abnormal signal intensity of olfactory bulbs in COVID-19 patients with OD were significantly higher [17].

Obstruction of OC is another possible underlying mechanism of OD. In a study on 305 patients with COVID-19 and OD that underwent various neuroimaging modalities, the most common finding was an obstruction in OC with normal morphology and T2/FLAIR signal intensity. Also, there were some cases of OB and OC atrophy in the late phase of COVID-19 patients, which can explain central mechanisms in the late stages of OD[41]. In this regard, one study showed that changes in OC structure are greater than those in OB which can be related to the different immune responses in OC [15].

Another explanation for OD is the disruption in sustentacular cells due to infection. These cells support olfactory sensory neurons by balancing the ions. Therefore, damage to the sustentacular cells can hamper sensory neurons signaling to OB. Consequently, OD will happen due to the partial lack of signal transmission [15].

Besides the olfactory structures, some adjacent areas in the brain are also reported to be affected by the virus, including the cingulate cortex, corpus callosum, and insula, which are parts of the olfactory network and can influence the senses of smell [42]. The Insula region plays an essential role in smell processing. The Corpus callosum connects two brain hemispheres, which contain olfactory cortices in the temporal lobes and helps the integration of sensory information. In terms of the cingulate cortex, an olfactory stimulation can trigger a functional pathway in the cingulate cortex to the olfactory system. One possible hypothesis is that systemic response to the virus might lead to inflammation and dysfunction of these areas, which are not direct targets of the virus [14].

### 4.2 Follow-up and recovery of patients with Olfactory dysfunction

According to case-control studies, OD is higher reported in COVID-19 than in other respiratory infections [3, 33]. The rate of OD in COVID-19 patients varied in different groups. However, different observational studies reported that 5.1% of hospitalized COVID-19 patients suffered from loss of smell [43, 44]. In other studies, the rate of OD in hospitalized and home-treated patients reached 73% and 85.6%, respectively [7, 45]. Moreover, nasal obstruction and rhinorrhea were found in 12.9%-40% and 18%-28.5% of OD patients, respectively. It was also reported that less severe COVID-19 was associated with a higher rate of anosmia [33].

Most COVID-19 patients with OD recovered during 7-10 days [33], while in some studies, it was reported to continue for up to 6 months [46, 47]. Remarkably, some studies did follow-up for OD patients, and only five studies concluded that OD symptoms persisted in less than 10% of patients for more than 20 days. Studies with longer follow-up times (117, 125 days) reported 88% and 84% recovery rates, respectively [48, 49]. Interestingly, a cohort study on 100 patients with moderate COVID-19 symptoms conducted follow-up for one year, and 48% of patients suffered from olfactory dysfunction. These patients experienced long term home-isolation, which can be associated with OD persistence. Additionally, some believe a higher viral load might lead to symptom persistence [50]. On the other hand, a systematic review of the 18 original studies reported that 33-36% of adult COVID-19 patients experienced incomplete OD recovery, and 3-13.9% suffered a lack of recovery during 12-39 days after symptoms onset [51]. Primarily, the presence of OD was associated with a mild course of COVID-19, which was indicated by lower inflammatory markers, ICU admission, oxygen therapy, hospitalization, intubation and mortality, and a decreased risk of pneumonia[19]. Although OD is more likely to persist in patients with moderate symptoms [52], anosmia was also reported in severe COVID-19 cases [32].

It was also recognized that OD symptoms can occur before other COVID-19 clinical manifestations and should be taken thoughtfully [30]. Regarding risk factors for OD, no definite consensus was reached on gender, and several studies displayed that gender is not a risk factor for OD [26]. Mild and severe COVID-19 patients with severe initial OD had inferior OD recovery and outcomes [26]. A meta-analysis reported that olfactory dysfunction is more likely to emerge in younger adults in high-income families [34]. Thus, age might be a risk factor for OD. It is believed that individuals aged<40 have better tissue resilience. Considering the results of a study demonstrating more prevalence of OD in patients older than 70, it seems rational to consider age as an influential factor [53]. However, there are controversial studies in that respect [54].

Smoking Cessation may accelerate OD recovery after COVID-19. However, researches on the effect of smoking are controversial[20]. Parosmia, distortion in the ability to smell known odorants, is also reported in COVID-19 patients and studies proved it can worsen OD. The reason behind this could be the emergence of immature neurons due to a COVID-19 induced decrease in repairing olfactory neurons [55]. A study reported that approximately half of COVID-19 patients suffered from parosmia for up to one and a half year after infection. Another predictive factor is trigeminal sensation. Some studies showed that the trigeminal neurons could be the targets of SARS-Cov-2. Besides, the trigeminal nerve itself is associated with the olfactory system. Thus, poor function of the trigeminal nerve might be a predictor of OD [56].

Dental flossing can reduce the risk of gingival inflammation. A significant higher rate of dental flossing was observed in the early-smell-recovery group compared to the late-recovery group. [28]. Platelet count was also mentioned as a predictor. SARS-Cov-2 particles can develop thrombosis through affecting on ACE-2 receptors on endothelium, which might occur in the olfactory system. A lower platelet count could reduce the chance of platelet aggregation, and thrombosis in the olfactory system [28].

### 4.3 Management of Olfactory dysfunction

#### 4.3.1 Non-pharmacological management

Based on previous studies almost one-third of patients with persistent symptoms of OD resolve spontaneously without any treatment. Because many patients also do not visit a physician, the percentage of spontaneous elimination of OD can be even more. The recovery rate might vary depending on the patient’s age, degree of OD, and duration [20].

Based on documents, including RCTs and meta-analysis, olfactory training is the treatment of choice for OD [20, 22]. OT therapy involves repeated exposure to odorants. It is believed to regenerate olfactory receptor neurons and improve olfactory information processing in patients who suffer olfactory loss related to different etiologies. The OT consist of using 4 odorants which should be smelled for five minutes twice daily and continued for at least 12 weeks [25]. Any odorant which can encourage patient’s compliance is recommended. One interesting study used a concurrent depiction of the scent coupled with smelling and showed that after 4 weeks, 64% of patient’s symptoms improved. Additionally, it was shown that earlier initiation of OT (<12 months) along with the long duration of it (up to 56weeks), was associated with olfactory dysfunction improvement [57].

Smoking cessation is another modality that improves olfactory function. Although it has been shown that smoking can worsen the severity of olfactory dysfunction, there are conflicting studies regarding its effectiveness on the loss of olfaction. A meta-analysis found that smoking is associated with an increased risk of olfactory dysfunction. In contrast, a retrospective study showed no association between smoking and olfactory function status [57].

Traditional Chinese acupuncture is a treatment for healing that has been used for centuries. A reanalyzed prospective study investigated its effect on PIOD and showed no improvement in PIOD patients [22].

#### 4.3.2 Pharmacological management

Pharmacological agents could also be useful for treating PIOD patients. Pharmacological drugs are recommended whenever the PIOD is sustained over 2 weeks. Some recent studies suggest that anti-inflammatory agents such as oral and intranasal corticosteroids can play a major role in respect of PIOD patients. Systemic or topical steroids can improve OD in chronic rhino sinusitis. Regarding intranasal corticosteroids (ICS), there are limited documents evaluating the effectiveness of their independent use. Studies mostly suggest ICS in combination with OT might be beneficial for patients. Also, short-term treatment is recommended because no difference was observed between short-term and long-term use of ICS. For symptoms lasting more than 2 weeks, patients can start using ICS. Regarding side effects, it depends on the amount of drug that can reach the olfactory cleft. The Kaiteki position is helpful in which an individual lay on one side, the head tilted, and the chin lifted at 20-40 degrees [57]. A meta-analysis conducted on OD recovery after four weeks post-COVID-19 treatment with topical steroids showed that ICS did not affect the rate of full recovery, while olfactory improvement within four weeks was statistically significantly more significant in the intervention group [23]. Thus, considering the low risk of these agents and limited side effects, a short-term trial as a complementary drug is recommended. Oral corticosteroids (OCS) have limited evidence of effectiveness and therefore, are not a part of routine clinical management. Interestingly, it was shown that patients with any cause of OD concluded that an initial short course of OCS can be practical while continuing them can have some side effects without more points. Nevertheless, before initiation, all risks and benefits should be evaluated for each patient [20].

It seems that an OCS trial can be helpful in patients who experience only olfactory symptoms of COVID-19. Similar to ICS, OCS, in combination with OT therapy, is proven to be more effective than OT therapy alone. In a non-randomized control trial with patients who suffered OD symptoms for more than one month, a combination of ICS and oral prednisolone for 40 days was significantly helpful. Thus, in refractory cases, it can make improvements. OCS therapy should be used based on the patient’s conditions, and a 3-4-day trial can be helpful initially [57]. Moreover, there are other non-steroid pharmacological agents which can be useful. It should be considered that no study specifically evaluates the efficacy of non-steroidal agents on the COVID-19 associated olfactory symptom. Therefore, the drugs are recommended based on their mechanisms of action and effectiveness on any causes of OD. Theophylline is proven to be effective in neural generation by influencing cell massages. One study on hyposmia showed approximately half of the patients benefited from theophylline [28]. Moreover, another study showed greater doses of theophylline could make more improvement in OD [58]. Sodium citrate can reduce calcium in the mucosa, which will be reduced the negative feedback and increase odorant sensitivity. A randomized clinical trial concluded that almost one-third of OD cases benefited from sodium citrate and improved their olfactory function[59]. Similarly, a cohort study showed significant improvement in OD in patients with PIOD after using sodium citrate [60]. However, some other studies have sparse outcomes.

Caroverine is a reversible antagonism of N-Methyl D-aspartate (NMDA) receptor and produced promising results. Although the exact underlying mechanism of caroverine on the neuroepithelium is not fully understood, an RCT study demonstrated that patients with hyposmia and anosmia showed improvement in odor identification following using 120 mg/daily of caroverine [61].

Alpha-lipoic acid (ALA) is a fatty acid that stimulates nerve growth factors expression, substance P and neuropeptide Y all of which have neuroprotective capabilities. In a prospective study, This also showed moderate olfactory improvement in 61% of its users [62]. Vitamin A can aid in the regeneration of olfactory neuro epithelium. Several studies showed it improves olfactory function after systematic treatment in patients with mixed causes of olfactory loss. Furthermore, a study found that using 10000 units Vit A combined with OT can improve olfactory dysfunction compared to OT alone [63].

Regarding zinc sulfate, some studies showed improvement in olfactory function compared with placebo and mometasone propionate. Zinc sulfate can potentially shorten the duration of OD symptoms without affecting the recovery rate. Considering the limitation of these studies, all of these therapeutic agents need more investigation to be a part of OD management. [20, 64]. Some studies investigated the effects of probiotics on COVID-19, and one showed that 14-day administration of L.lactis W136 intranasal spray could reduce the olfactory dysfunction rate in COVID-19 patients. [65].

Topical intranasal insulin also showed improvement in this regard. A clinical trial showed that intranasal administration of 40 IU insulin in approximately 60% of patients improved odor threshold and increased the detection of odors [66].

## Conclusion

Based on documents, the COVID-19 virus can cause abnormalities in olfactory system structures. Opacification of olfactory cleft, olfactory bulb deformation and degeneration in the delayed phase, T2 and FLAIR hyper-intensity, and decreased PET metabolism in the olfactory cortex are neuroimaging findings of COVID-19 with OD. Additionally, the virus or inflammation can affect some brain regions associated with the olfactory system, including the cingulate cortex, corpus callosum, and insula. Various reports showed that a high proportion of infected patients had olfactory system-associated symptoms and the recovery rate varies based on the patient’s condition, such as age. Generally, younger patients with mild clinical manifestation are more likely to experience OD, especially the persistent type. In many cases, OD symptoms resolve up to 2 weeks after their initiation; these symptoms can persist in a low percentage of patients. As predictors of OD recovery, many factors, such as sex, age, and severity of COVID-19, have been discussed in numerous studies with varying results.

Regarding management, the most effective was olfactory training (OT), a non-pharmacological treatment that should continue for at least three months. Some research indicates that topical corticosteroids improve odor threshold and recognition, while others indicate that the effect of corticosteroids on olfactory function and the adverse effects of steroids are yet unknown. Oral corticosteroids can be helpful in limited situations, and this decision depends on each patient’s condition. Moreover, some non-steroid agents can be helpful due to their effectiveness on neuro generation, among which vitamin A had promising effects. Other drugs such as zinc sulfate, theophylline, and probiotics, produced conflicting results. Most research suggests olfactory training as the initial treatment for post-infectious olfactory impairment.

## Supporting information

Search Strategies and Quality Assessment Table

## Data Availability

All data produced in the present study are available upon reasonable request to the authors.

---

**Table 1.**
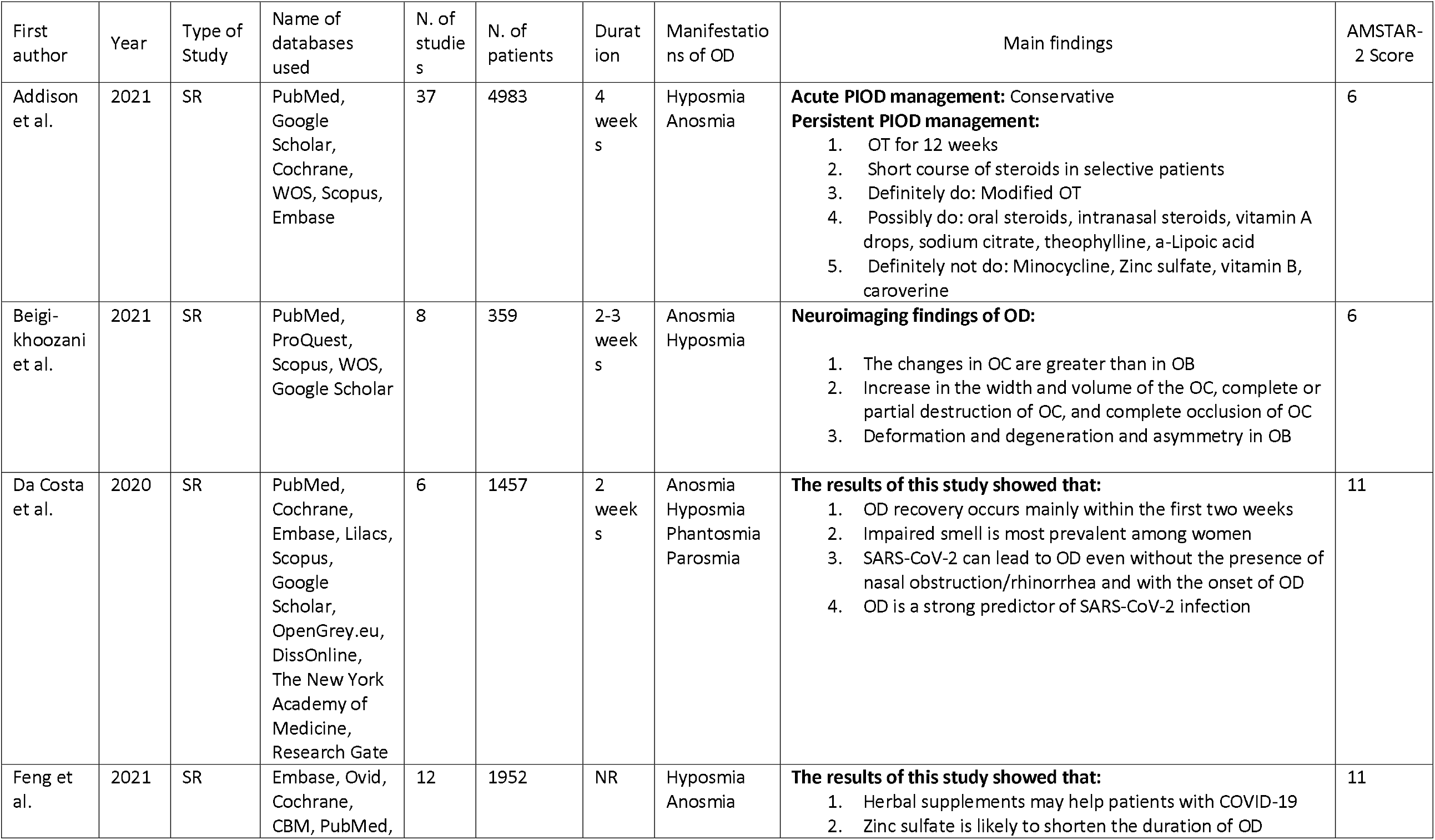
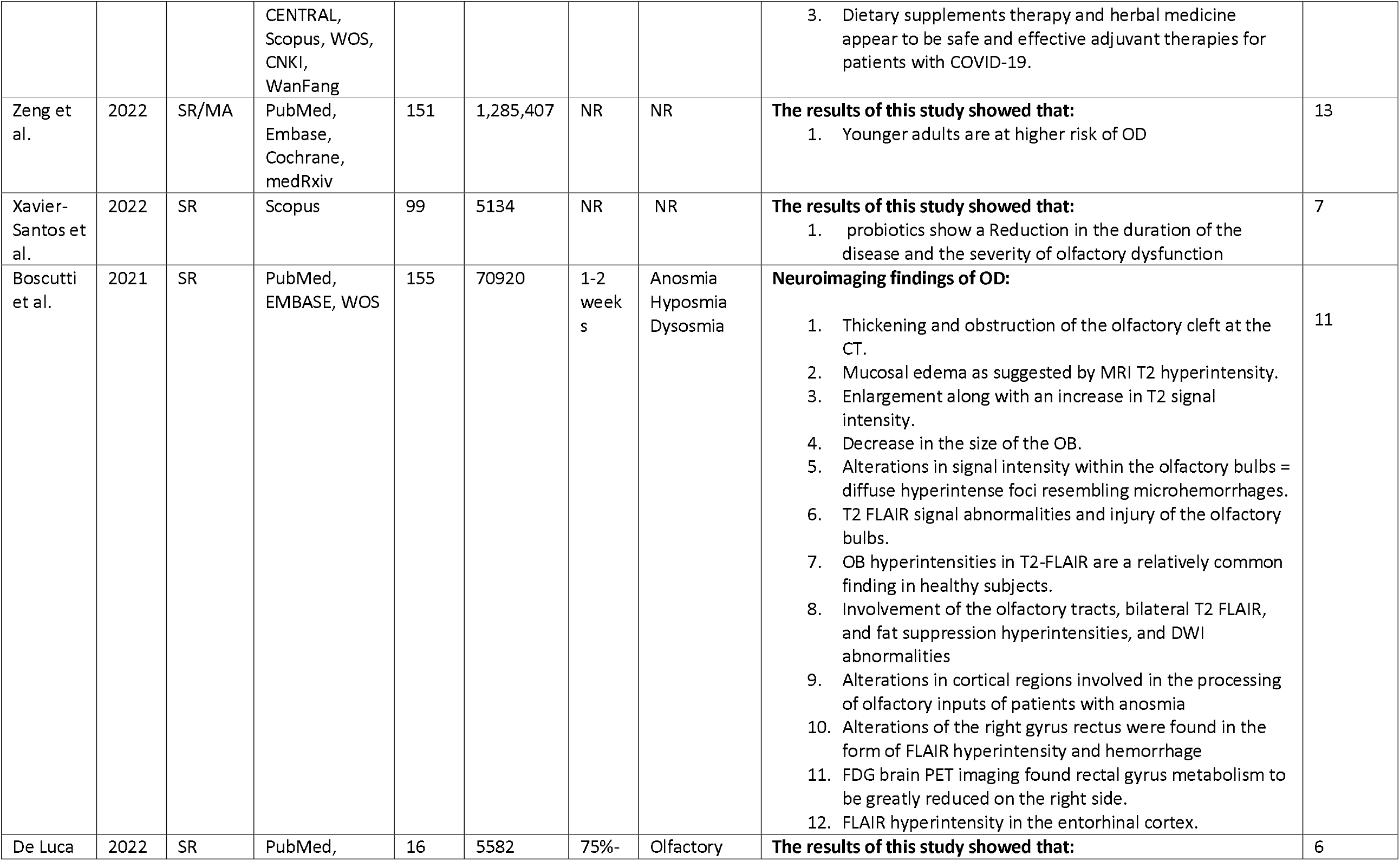
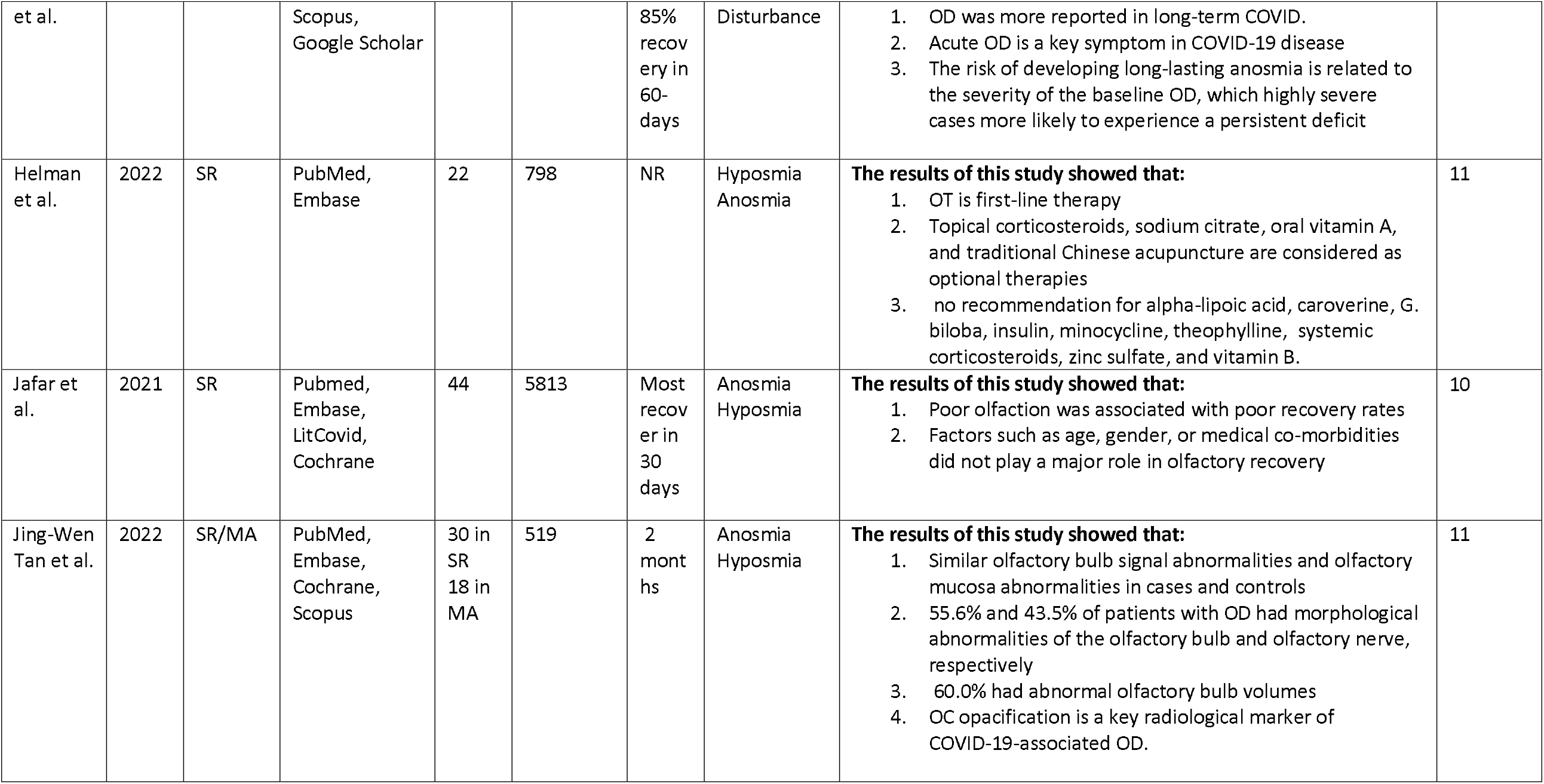
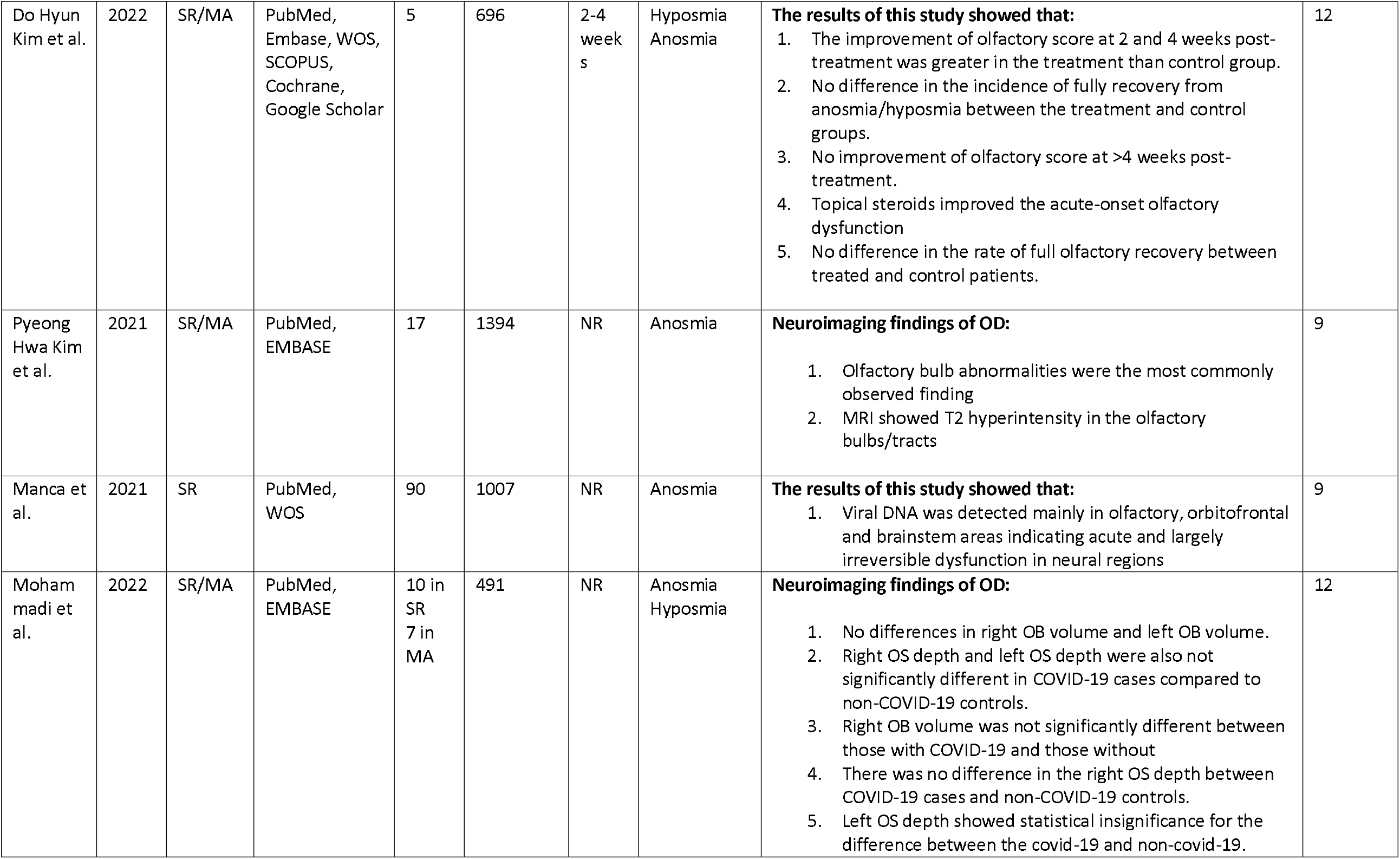
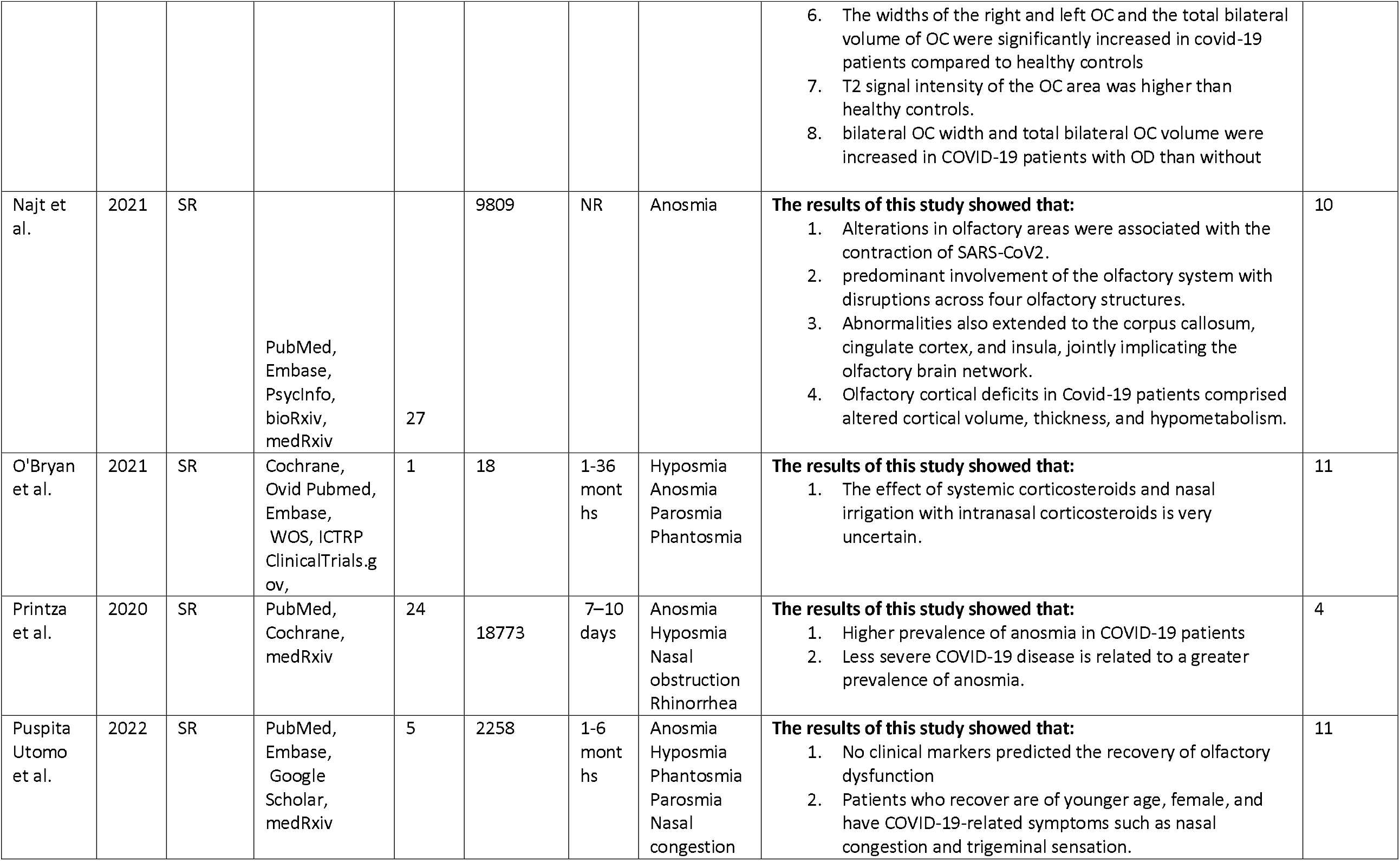
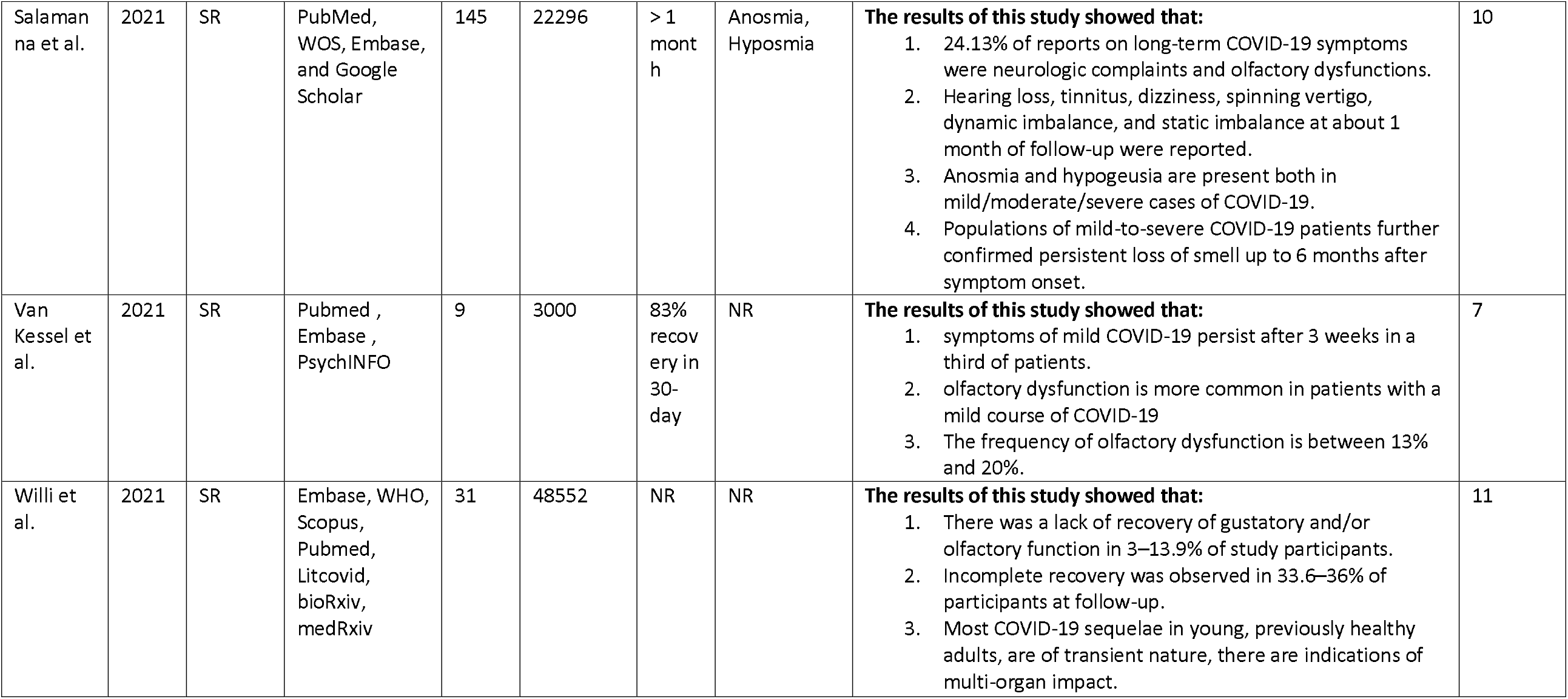
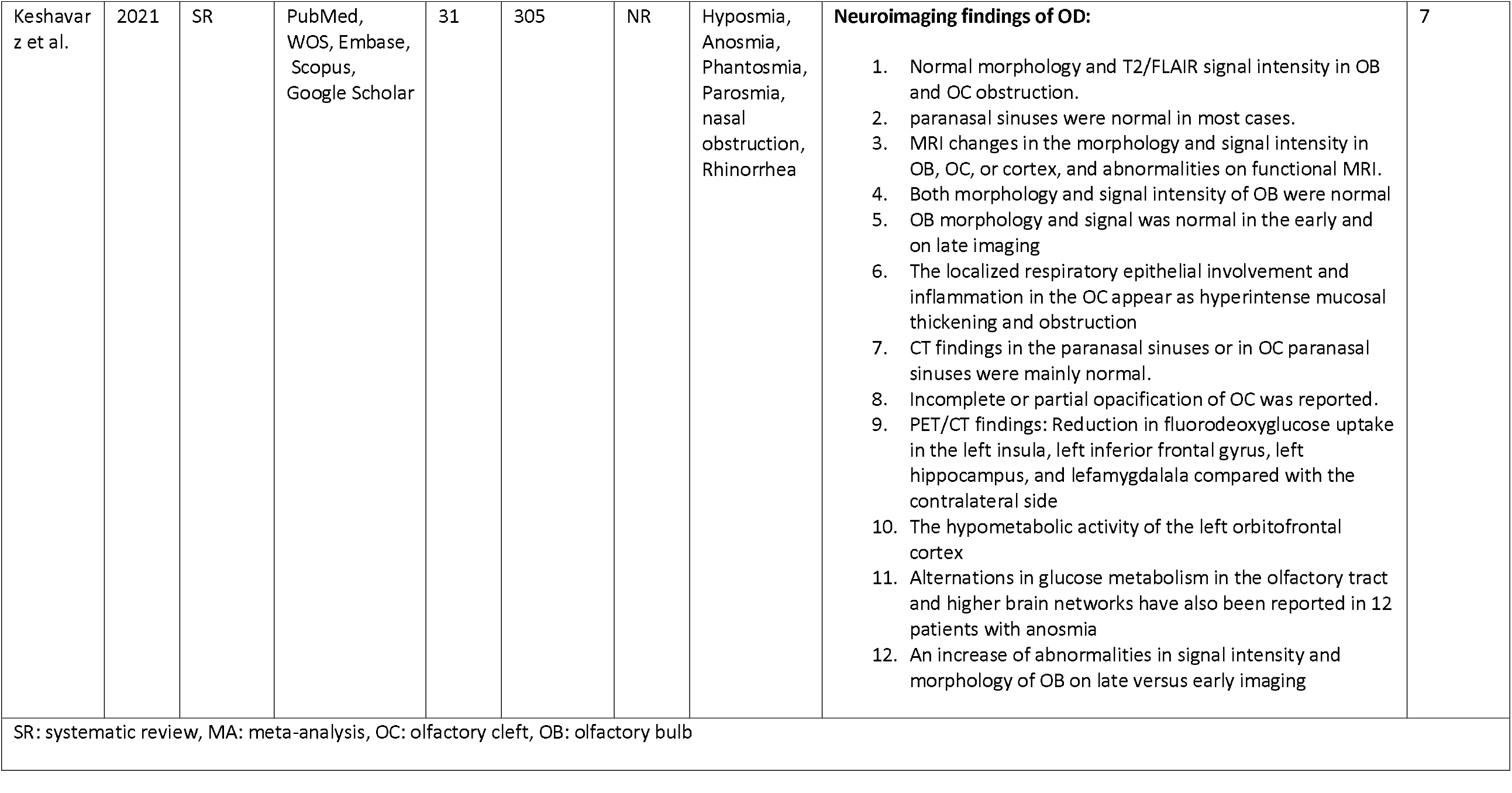
Data extraction table.

## References

1. Mogharab, V., et al., Global burden of the COVID-19 associated patient-related delay in emergency healthcare: a panel of systematic review and meta-analyses. Globalization and Health, 2022. 18(1): p. 58.

2. Zaim, S., et al., COVID-19 and Multiorgan Response. Current problems in cardiology, 2020. 45(8): p. 100618–100618.

3. Moein, S.T., et al., Smell dysfunction: a biomarker for COVID-19. Int Forum Allergy Rhinol, 2020. 10(8): p. 944–950.

4. Saniasiaya, J. and N. Prepageran, Impact of olfactory dysfunction on quality of life in coronavirus disease 2019 patients: a systematic review. J Laryngol Otol, 2021. 135(11): p. 947–952.

5. Pekala, K., R.K. Chandra, and J.H. Turner, Efficacy of olfactory training in patients with olfactory loss: a systematic review and meta-analysis. Int Forum Allergy Rhinol, 2016. 6(3): p. 299–307.

6. Le Bon, S.D., et al., Efficacy and safety of oral corticosteroids and olfactory training in the management of COVID-19-related loss of smell. Eur Arch Otorhinolaryngol, 2021. 278(8): p. 3113–3117.

7. Lechien, J.R., et al., Olfactory and gustatory dysfunctions as a clinical presentation of mild-to-moderate forms of the coronavirus disease (COVID-19): a multicenter European study. Eur Arch Otorhinolaryngol, 2020. 277(8): p. 2251–2261.

8. Carrillo-Larco, R.M. and C. Altez-Fernandez, Anosmia and dysgeusia in COVID-19: A systematic review. Wellcome Open Res, 2020. 5: p. 94.

9. Mendelson, M., et al., Long-COVID: An evolving problem with an extensive impact. S Afr Med J, 2020. 111(1): p. 10–12.

10. Liberati, A., et al., The PRISMA statement for reporting systematic reviews and meta-analyses of studies that evaluate health care interventions: explanation and elaboration. PLoS Med, 2009. 6(7): p. e1000100.

11. Aromataris, E., et al., Summarizing systematic reviews: methodological development, conduct and reporting of an umbrella review approach. Int J Evid Based Healthc, 2015. 13(3): p. 132–40.

12. Shea, B.J., et al., AMSTAR 2: a critical appraisal tool for systematic reviews that include randomised or non-randomised studies of healthcare interventions, or both. BMJ, 2017. 358: p. j4008.

13. Keshavarz, P., et al., A Systematic Review of Imaging Studies in Olfactory Dysfunction Secondary to COVID-19. Acad Radiol, 2021. 28(11): p. 1530–1540.

14. Najt, P., H.L. Richards, and D.G. Fortune, Brain imaging in patients with COVID-19: A systematic review. Brain Behav Immun Health, 2021. 16: p. 100290.

15. Beigi-Khoozani, A., A. Merajikhah, and M. Soleimani, Magnetic Resonance Imaging Findings of Olfactory Bulb in Anosmic Patients with COVID-19: A Systematic Review. Chin Med Sci J, 2022. 37(1): p. 23–30.

16. Kim, P.H., et al., Neuroimaging Findings in Patients with COVID-19: A Systematic Review and Meta-Analysis. Korean J Radiol, 2021. 22(11): p. 1875–1885.

17. Tan, C.J., et al., Neuroradiological Basis of COVID-19 Olfactory Dysfunction: A Systematic Review and Meta-Analysis. Laryngoscope, 2022. 132(6): p. 1260–1274.

18. Mohammadi, S., et al., Olfactory system measurements in COVID-19: a systematic review and meta-analysis. Neuroradiology, 2022: p. 1–15.

19. Boscutti, A., et al., Olfactory and gustatory dysfunctions in SARS-CoV-2 infection: A systematic review. Brain Behav Immun Health, 2021. 15: p. 100268.

20. Addison, A.B., et al., Clinical Olfactory Working Group consensus statement on the treatment of postinfectious olfactory dysfunction. J Allergy Clin Immunol, 2021. 147(5): p. 1704–1719.

21. O’Byrne, L., et al., Interventions for the treatment of persistent post-COVID-19 olfactory dysfunction. Cochrane Database Syst Rev, 2021. 7(7): p. Cd013876.

22. Helman, S.N., et al., Treatment strategies for postviral olfactory dysfunction: A systematic review. Allergy Asthma Proc, 2022. 43(2): p. 96–105.

23. Kim, D.H., et al., Efficacy of topical steroids for the treatment of olfactory disorders caused by COVID-19: A systematic review and meta-analysis. Clin Otolaryngol, 2022. 47(4): p. 509–515.

24. Dhar, S., J. Seth, and D. Parikh, Systemic side-effects of topical corticosteroids. Indian journal of dermatology, 2014. 59(5): p. 460–464.

25. Hummel, T., et al., Effects of olfactory training in patients with olfactory loss. The Laryngoscope, 2009. 119(3): p. 496–499.

26. Jafar, A., et al., Olfactory recovery following infection with COVID-19: A systematic review. PLoS One, 2021. 16(11): p. e0259321.

27. Willi, S., et al., COVID-19 sequelae in adults aged less than 50 years: A systematic review. Travel Med Infect Dis, 2021. 40: p. 101995.

28. Utomo, N.P. and A.D. Iswarini, Impaired olfaction post-coronavirus disease 2019: a systematic review of smell recovery predictive factors. The Egyptian Journal of Otolaryngology, 2022. 38(1): p. 80.

29. De Luca, P., et al., Long COVID, audiovestibular symptoms and persistent chemosensory dysfunction: a systematic review of the current evidence. Acta Otorhinolaryngol Ital, 2022. 42(Suppl. 1): p. S87–s93.

30. Costa, K., et al., Olfactory and taste disorders in COVID-19: a systematic review. Braz J Otorhinolaryngol, 2020. 86(6): p. 781–792.

31. van Kessel, S.A.M., et al., Post-acute and long-COVID-19 symptoms in patients with mild diseases: a systematic review. Fam Pract, 2022. 39(1): p. 159–167.

32. Salamanna, F., et al., Post-COVID-19 Syndrome: The Persistent Symptoms at the Post-viral Stage of the Disease. A Systematic Review of the Current Data. Front Med (Lausanne), 2021. 8: p. 653516.

33. Printza, A. and J. Constantinidis, The role of self-reported smell and taste disorders in suspected COVIDIZI19. Eur Arch Otorhinolaryngol, 2020. 277(9): p. 2625–2630.

34. Zeng, N., et al., A systematic review and meta-analysis of long term physical and mental sequelae of COVID-19 pandemic: call for research priority and action. Molecular Psychiatry, 2022.

35. Pazoki, M., et al., Association of clinical characteristics, antidiabetic and cardiovascular agents with diabetes mellitus and COVID-19: a 7-month follow-up cohort study. J Diabetes Metab Disord, 2021. 20(2): p. 1545–1555.

36. Pang, K.W., et al., Frequency and Clinical Utility of Olfactory Dysfunction in COVID-19: a Systematic Review and Meta-analysis. Current Allergy and Asthma Reports, 2020. 20(12): p. 76.

37. Bilinska, K. and R. Butowt, Anosmia in COVID-19: A Bumpy Road to Establishing a Cellular Mechanism. ACS Chemical Neuroscience, 2020. 11(15): p. 2152–2155.

38. Whitcroft, K.L. and T. Hummel, Olfactory Dysfunction in COVID-19: Diagnosis and Management. JAMA, 2020. 323(24): p. 2512–2514.

39. Manca, R., et al., Heterogeneity in Regional Damage Detected by Neuroimaging and Neuropathological Studies in Older Adults With COVID-19: A Cognitive-Neuroscience Systematic Review to Inform the Long-Term Impact of the Virus on Neurocognitive Trajectories. Front Aging Neurosci, 2021. 13: p. 646908.

40. Mohammadi, S., et al., Olfactory system measurements in COVID-19: a systematic review and meta-analysis. Neuroradiology, 2022.

41. Keshavarz, P., et al., A Systematic Review of Imaging Studies in Olfactory Dysfunction Secondary to COVID-19. Academic Radiology, 2021. 28(11): p. 1530–1540.

42. Chung, T.W., et al., Neurosensory Rehabilitation and Olfactory Network Recovery in Covid-19-related Olfactory Dysfunction. Brain Sci, 2021. 11(6).

43. Beltrán-Corbellini, Á., et al., Acute-onset smell and taste disorders in the context of COVID-19: a pilot multicentre polymerase chain reaction based case-control study. Eur J Neurol, 2020. 27(9): p. 1738–1741.

44. Mao, L., et al., Neurologic Manifestations of Hospitalized Patients With Coronavirus Disease 2019 in Wuhan, China. JAMA Neurol, 2020. 77(6): p. 683–690.

45. Vaira, L.A., et al., Objective evaluation of anosmia and ageusia in COVID-19 patients: Single-center experience on 72 cases. Head Neck, 2020. 42(6): p. 1252–1258.

46. Petrocelli, M., et al., Six-month smell and taste recovery rates in coronavirus disease 2019 patients: a prospective psychophysical study. The Journal of Laryngology & Otology, 2021. 135(5): p. 436–441.

47. Riestra-Ayora, J., et al., Long-term follow-up of olfactory and gustatory dysfunction in COVID-19: 6 months case–control study of health workers. European Archives of Oto-Rhino-Laryngology, 2021. 278(12): p. 4831–4837.

48. Li, J., et al., Olfactory Dysfunction in Recovered Coronavirus Disease 2019 (COVID-19) Patients. Movement Disorders, 2020. 35(7): p. 1100–1101.

49. Stavem, K., et al., Persistent symptoms 1.5–6 months after COVID-19 in non-hospitalised subjects: a population-based cohort study. Thorax, 2021. 76(4): p. 405–407.

50. De Luca, P., et al., Long COVID, audiovestibular symptoms and persistent chemosensory dysfunction: a systematic review of the current evidence. Acta otorhinolaryngologica Italica : organo ufficiale della Societa italiana di otorinolaringologia e chirurgia cervico-facciale, 2022. 42(Suppl. 1): p. S87–S93.

51. Willi, S., et al., COVID-19 sequelae in adults aged less than 50 years: A systematic review. Travel Medicine and Infectious Disease, 2021. 40: p. 101995.

52. van Kessel, S.A.M., et al., Post-acute and long-COVID-19 symptoms in patients with mild diseases: a systematic review. Family Practice, 2022. 39(1): p. 159–167.

53. Pellegrino, R., et al., Prevalence and correlates of parosmia and phantosmia among smell disorders. Chem Senses, 2021. 46.

54. Nguyen, N.N., et al., Long-term persistence of olfactory and gustatory disorders in COVID-19 patients. Clinical microbiology and infection : the official publication of the European Society of Clinical Microbiology and Infectious Diseases, 2021. 27(6): p. 931–932.

55. Leopold, D.A., T.A. Loehrl, and J.E. Schwob, Long-term follow-up of surgically treated phantosmia. Arch Otolaryngol Head Neck Surg, 2002. 128(6): p. 642–7.

56. Desforges, M., et al., Human Coronaviruses and Other Respiratory Viruses: Underestimated Opportunistic Pathogens of the Central Nervous System? Viruses, 2019. 12(1).

57. Wu, T.J., A.C. Yu, and J.T. Lee, Management of post-COVID-19 olfactory dysfunction. Curr Treat Options Allergy, 2022. 9(1): p. 1–18.

58. Henkin, R.I., I. Velicu, and L. Schmidt, An open-label controlled trial of theophylline for treatment of patients with hyposmia. Am J Med Sci, 2009. 337(6): p. 396–406.

59. Philpott, C.M., et al., A randomised controlled trial of sodium citrate spray for non-conductive olfactory disorders. Clinical Otolaryngology, 2017. 42(6): p. 1295–1302.

60. Whitcroft, K.L., et al., Intranasal sodium citrate solution improves olfaction in post-viral hyposmia. Rhinology, 2016. 54(4): p. 368–374.

61. Quint, C., et al., The quinoxaline derivative caroverine in the treatment of sensorineural smell disorders: A proof-of-concept study. Acta Oto-Laryngologica, 2002. 122(8): p. 877–881.

62. Hummel, T., S. Heilmann, and K.B. Hüttenbriuk, Lipoic acid in the treatment of smell dysfunction following viral infection of the upper respiratory tract. Laryngoscope, 2002. 112(11): p. 2076–80.

63. Hummel, T., et al., Intranasal vitamin A is beneficial in post-infectious olfactory loss. Eur Arch Otorhinolaryngol, 2017. 274(7): p. 2819–2825.

64. Feng, Z., et al., Dietary supplements and herbal medicine for COVID-19: A systematic review of randomized control trials. Clinical nutrition ESPEN, 2021. 44: p. 50–60.

65. Xavier-Santos, D., et al., Evidences and perspectives of the use of probiotics, prebiotics, synbiotics, and postbiotics as adjuvants for prevention and treatment of COVID-19: A bibliometric analysis and systematic review. Trends in Food Science & Technology, 2022. 120: p. 174–192.

66. Schöpf, V., et al., Intranasal insulin influences the olfactory performance of patients with smell loss, dependent on the body mass index: A pilot study. Rhinology, 2015. 53(4): p. 371–8.

