## Supplementary material for "Olfactory Dysfunction in the COVID-19 Era: An Umbrella Review Focused on Neuroimaging, Management, and Follow-up": Search Strategies and Quality Assessment Table

**Table S1:** Search strategies for various databases.

| Database | Search strategy | Number |
| --- | --- | --- |
| PubMed | (Olfactory[tiab]) AND (“COVID-19”[mesh] OR “SARS-CoV-2”[mesh] OR COVID-19[tiab] OR SARS-CoV-2[tiab] OR coronavirus disease 2019[tiab] OR severe acute respiratory syndrome coronavirus 2[tiab]) AND (systematic review[tiab]) | 89 |
| Embase | (‘Olfactory’:ab,ti) AND (’coronavirus disease 2019’/exp OR ’severe acute respiratory syndrome coronavirus 2’/exp OR ’COVID-19’:ab,ti OR ’SARS-CoV-2’:ab,ti OR ’coronavirus disease 2019’:ab,ti OR ’severe acute respiratory syndrome coronavirus 2’:ab,ti) AND (‘systematic review’:ab,ti) | 85 |
| Scopus | (TITLE-ABS (“Olfactory”)) AND (TITLE-ABS (“COVID-19” OR “SARS-CoV-2” OR “coronavirus disease 2019” OR “severe acute respiratory syndrome coronavirus 2”) AND (TITLE-ABS (“systematic review”)) | 87 |
| Web of Science | (TS= (“Olfactory”)) AND (TS= (“COVID-19” OR “SARS-CoV-2” OR “coronavirus disease 2019” OR “severe acute respiratory syndrome coronavirus 2”)) AND (TS= (“systematic review”)) | 83 |

mesh, Medical Subject Headings.

**Table S2:** Quality assessment table.

| Articles | 1 | 2 | 3 | 4 | 5 | 6 | 7 | 8 | 9 | 10 | 11 | 12 | | 13 | 14 | 15 | 16 | AMSTAR-2 Score |
| --- | --- | --- | --- | --- | --- | --- | --- | --- | --- | --- | --- | --- | --- | --- | --- | --- | --- | --- |
| Printza et al. | No | No | Yes | Yes | No | No | Partial Yes | Partial Yes | No | No | No meta-analysis conducted | | No meta-analysis conducted | No | No | No meta-analysis conducted | No | 4 |
| Boscutti et al. | Yes | Partial Yes | Yes | Yes | Yes | No | Yes | Yes | Yes | Yes | No meta-analysis conducted | No meta-analysis conducted | | Yes | Yes | No meta-analysis conducted | No | 11 |
| da Costa et al. | Yes | Yes | Yes | Yes | Yes | Yes | Yes | Yes | Yes | No | No meta-analysis conducted | No meta-analysis conducted | | Yes | Yes | No meta-analysis conducted | No | 11 |
| Jafar et al. | No | Yes | Yes | Partial Yes | Yes | Yes | Yes | Yes | Yes | No | No meta-analysis conducted | No meta-analysis conducted | | Yes | Yes | No meta-analysis conducted | No | 10 |
| Utomo et al. | Yes | Yes | Yes | Partial Yes | Yes | Yes | Yes | Yes | Yes | No | No meta-analysis conducted | No meta-analysis conducted | | Yes | Yes | No meta-analysis conducted | No | 11 |
| Mohammadi et al. | No | Yes | Yes | Partial Yes | Yes | Yes | Partial Yes | Yes | Yes | No | Yes | Yes | | Yes | Yes | No | No | 12 |
| Manca et al. | Yes | No | Yes | Yes | Yes | Yes | Yes | Yes | No | Yes | No meta-analysis conducted | No meta-analysis conducted | | No | Yes | No meta-analysis conducted | No | 9 |
| Hwa Kim et al. | Yes | No | Yes | Yes | Yes | Yes | Yes | Yes | No | No | Yes | No | | No | Yes | No | No | 9 |
| Jing-Wen Tan et al. | Yes | Yes | Yes | Yes | Yes | Yes | Partial Yes | Yes | Yes | No | Yes | No | | No | Yes | No | No | 11 |
| Keshavarz et al. | No | No | Yes | Yes | Yes | Yes | Partial Yes | Yes | No | No | No meta-analysis conducted | No meta-analysis conducted | | No | Yes | No meta-analysis conducted | No | 7 |
| Beigi-khoozani et al. | No | No | Yes | Partial Yes | No | Yes | Partial Yes | Partial Yes | No | No | No meta-analysis conducted | No meta-analysis conducted | | No | Yes | No meta-analysis conducted | No | 6 |
| Najt et al. | Yes | Yes | Yes | Yes | Yes | Yes | Yes | Yes | Yes | No | No meta-analysis conducted | No meta-analysis conducted | | No | Yes | No meta-analysis conducted | No | 10 |
| Addison et al. | Yes | No | Yes | Yes | No | No | Partial Yes | Yes | Yes | No | No meta-analysis conducted | No meta-analysis conducted | | No | No | No meta-analysis conducted | No | 6 |
| O'Byrne et al. | Yes | Partial Yes | Yes | Yes | Yes | Yes | Yes | Yes | Yes | No | No meta-analysis conducted | No meta-analysis conducted | | Yes | No | Yes | No | 11 |
| Hyun Kim et al. | Yes | Yes | Yes | Yes | Yes | Yes | Partial Yes | Yes | Yes | Yes | Yes | No | | No | Yes | No | No | 12 |
| Feng et al. | Yes | No | Yes | Partial Yes | Yes | Yes | Yes | Yes | Yes | Yes | No meta-analysis conducted | No meta-analysis conducted | | Yes | Yes | No meta-analysis conducted | No | 11 |
| Xavier-Santos et al. | Yes | No | Yes | Yes | Yes | No | No | Yes | No | Yes | No meta-analysis conducted | No meta-analysis conducted | | No | Yes | No meta-analysis conducted | No | 7 |
| Helman et al. | Yes | Yes | Yes | Yes | Yes | Yes | Yes | Yes | Yes | No | No meta-analysis conducted | No meta-analysis conducted | | Yes | Yes | No meta-analysis conducted | No | 11 |
| Zeng et al. | No | Yes | Yes | Partial Yes | Yes | Yes | Yes | Yes | Yes | Yes | Yes | Yes | | Yes | Yes | No | No | 13 |
| De Luca et al. | No | No | No | Yes | Yes | Yes | Yes | Yes | No | No | No meta-analysis conducted | No meta-analysis conducted | | No | Yes | No meta-analysis conducted | No | 6 |
| Van Kessel et al. | No | No | No | Partial Yes | Yes | Yes | Yes | Yes | Yes | No | No meta-analysis conducted | No meta-analysis conducted | | No | Yes | No meta-analysis conducted | No | 7 |
| Salamanna et al. | Yes | No | Yes | Yes | No | Yes | Yes | Yes | Yes | Yes | No meta-analysis conducted | No meta-analysis conducted | | Yes | Yes | No meta-analysis conducted | No | 10 |
| Willi et al. | No | Yes | Yes | Yes | Yes | Yes | Yes | Yes | Yes | Yes | No meta-analysis conducted | No meta-analysis conducted | | Yes | Yes | No meta-analysis conducted | No | 11 |

Quality assessment table by AMSTAR-2 tool. Items from 1 to 16 are as follows:

1. Did the research questions and inclusion criteria for the review include the components of PICO?
2. Did the report of the review contain an explicit statement that the review methods were established prior to conduct of the review and did the report justify any significant deviations from the protocol?
3. Did the review authors explain their selection of the study designs for inclusion in the review?
4. Did the review authors use a comprehensive literature search strategy?
5. Did the review authors perform study selection in duplicate?
6. Did the review authors perform data extraction in duplicate?
7. Did the review authors provide a list of excluded studies and justify the exclusions?
8. Did the review authors describe the included studies in adequate detail?
9. Did the review authors use a satisfactory technique for assessing the risk of bias (RoB) in individual studies that were included in the review?
10. Did the review authors report on the sources of funding for the studies included in the review?
11. If meta-analysis was justified did the review authors use appropriate methods for statistical combination of results?
12. If meta-analysis was performed did the review authors assess the potential impact of RoB in individual studies on the results of the meta-analysis or other evidence synthesis?
13. Did the review authors account for RoB in individual studies when interpreting/ discussing the results of the review?
14. Did the review authors provide a satisfactory explanation for, and discussion of, any heterogeneity observed in the results of the review?
15. If they performed quantitative synthesis did the review authors carry out an adequate investigation of publication bias (small study bias) and discuss its likely impact on the results of the review?
16. Did the review authors report any potential sources of conflict of interest, including any funding they received for conducting the review?
